# Red blood cells injuries and hypersegmented neutrophils in COVID-19 peripheral blood film

**DOI:** 10.1101/2020.07.24.20160101

**Authors:** Nordine Lakhdari, Boualem Tabet, Lila Boudraham, Malha Laoussati, Sofiane Aissanou, Lamia Beddou, Sihem Bensalem, Yuva Bellik, Lamine Bournine, Sofiane Fatmi, Mokrane Iguer-Ouada

**Affiliations:** Department of Hematology, University Hospital Center, Abderrahmane Mira University 06000 Bejaia, Algeria; Department of Internal Medicine, University Hospital Center, Abderrahmane Mira University 06000 Bejaia, Algeria; AME Laboratory, Faculty of Nature and Life Sciences, Abderrahmane Mira University 06000 Bejaia, Algeria

**Keywords:** Red blood cells, hypersegmented neutrophils, COVID-19, inflammation, oxidative stress

## Abstract

In the current investigation, peripheral blood films of 15 COVID-19 patients (44.78±16.55 years), proven by computed tomographic imaging and RT-PCR for coronavirus SARS-CoV-2, were analyzed at the moment of hospital admission. Blood tests showed raised inflammatory markers (C-reactive protein 58.2±61.2 mg/L) with normal values for hemoglobin (126.2±2.6 g/L), WBC (6.8±18.74 10^9^/L) RBC (4.55±0.99 10^12^/L) platelets (262.4±141.8, 10^9^/L) MCV (79.84±8.2 fL) MCH (28±3.31 pg) and MCHC (350.3±1.15 g/L). The results revealed the presence of hypersegmented neutrophils in 66.66%% of the patients. The percentages of neutrophils with 4 and 5 lobes were 46.25 ± 4.83% and 31.5 ± 14.84%, respectively. Three major red blood cells morphological alteration were observed: (1) erythrocytes in “rouleaux” formation represented by linear erythrocytes aggregation, (2) spherocytes with the disappearance of the usual biconcave disk, and (3) echinocytes showing spiky projections. Apparent reorganization of hemoglobin is found in the majority of the analyzed erythrocytes. Rouleaux formation is observed in 33.33% of patients and spherocytes and echinocytes are present at variable levels in the all analyzed patients. The current results revealed erythrocytes injuries in COVID-19 peripheral blood, in association with hypersegmented neutrophils, alterations that could be involved in the respiratory syndrome.

## Cases reports

Since the first patient reports in Wuhan, China, COVID-19 pandemic has spread worldwide with more than 15 million cases and over 600 000 deaths on late July 2020. SARS-CoV-2 (severe acute respiratory syndrome coronavirus 2) enters epithelial cells by interacting with the human angiotensin-converting enzyme receptor (ACE2) [1] causing acute respiratory syndrome with intense cytokine storm, hyperinflammatory reaction and multi-organ failure [2]. There is evidence that SARS-CoV-2 attacks blood vessels with injuries in endothelial cells, widespread thrombosis and micro angiopathy [3]. Peripheral blood films revealed neutrophil morphological abnormalities, immature granulocytes [4] and atypical lymphocytes[5] [6]. However, erythrocytes injuries are rarely reported in the literature. In two respective case reports, a mild leuco-erythroblastic picture with mild anisocytosis, rare dacrocytes [7] and red blood cells (RBCs) agglutination are reported [6].

In the current investigation, peripheral blood films of 15 COVID-19 patients (44.78±16.55 years), proven by computed tomographic imaging and RT-PCR for coronavirus SARS-CoV-2, were analyzed at the moment of hospital admission when antiviral and anti-inflammatory treatment was not yet administered. Blood tests showed raised inflammatory markers (C-reactive protein 58.2±61.2 mg/L) with normal values for hemoglobin (126.2±2.6 g/L), WBC (6.8±18.74 10^9^/L) RBC (4.55±0.99 10^12^/L) platelets (262.4±141.8, 10^9^/L) MCV (79.84±8.2 fL) MCH (28±3.31 pg) and MCHC (350.3±1.15 g/L).

Cells were analyzed in the area of a smear where RBCs are not overlapping and at the end of the smear. The results revealed the presence of hypersegmented neutrophils in 66.66%% of the patients. The percentages of neutrophils with 4 and 5 lobes were 46.25 ± 4.83% and 31.5 ± 14.84% respectively, demonstrating a marked shift to the right of the Arneth score. Neutrophil hypersegmentation is reported previously in cases of inflammation, acute respiratory distress syndrome [8] and severe viral respiratory infection [9].

The peripheral blood smears showed three major RBCs morphological alteration: (1) erythrocytes in “rouleaux” formation represented by linear erythrocytes aggregation, (2) spherocytes with the disappearance of the usual biconcave disk, appearing smaller than normal erythrocytes, and (3) echinocytes showing spiky projections. Apparent reorganization of hemoglobin is found in the majority of the analyzed red blood cells. Rouleaux formation is observed in 33.33% of patients. Spherocytes and echinocytes are present at variable levels in the all analyzed patients.

Reports showed that echinocytes and spherocytes are significantly correlated with oxidative stress and inflammation markers [10]. Particularly, under oxidative stress normal red blood cells transform into transient echinocytes and then to spherocytes [11]. Fibrinogen and immunoglobulin neutralize the membrane negative charge and allow the erythrocytes to aggregate in disease states with inflammatory reaction and oxidative stress [12]. Additionally, high erythrocyte sedimentation rate (ESR) is attributed to rouleaux formation and to other red blood cells agglutinations which compromises consequently tissue oxygenation [12]. Erythrocytes aggregation leads to vascular thrombosis and ischemia with multi-organ failure, alterations similar to those observed in COVID-19. Additionally, in COVID-19, SARS-CoV-2 by attaching to ACE 2 could increase angiotensin II and reduce angiotensin 1-7 leading to overexpression of oxidative stress [13]. This in turn, will enhance inflammation and cause endothelial cells and erythrocytes dysfunction.

In summary, the current findings pointed out the presence of abnormal red blood cells and hypersegmented neutrophils in peripheral blood coinciding with hospital admission of COVID-19 patients. Such abnormalities could be related to the cytokine storm and hyperinflammation with an overexpression of oxidative stress. Our observation calls for further studies with more patients, including different clinical phases.

**Figure 1.**
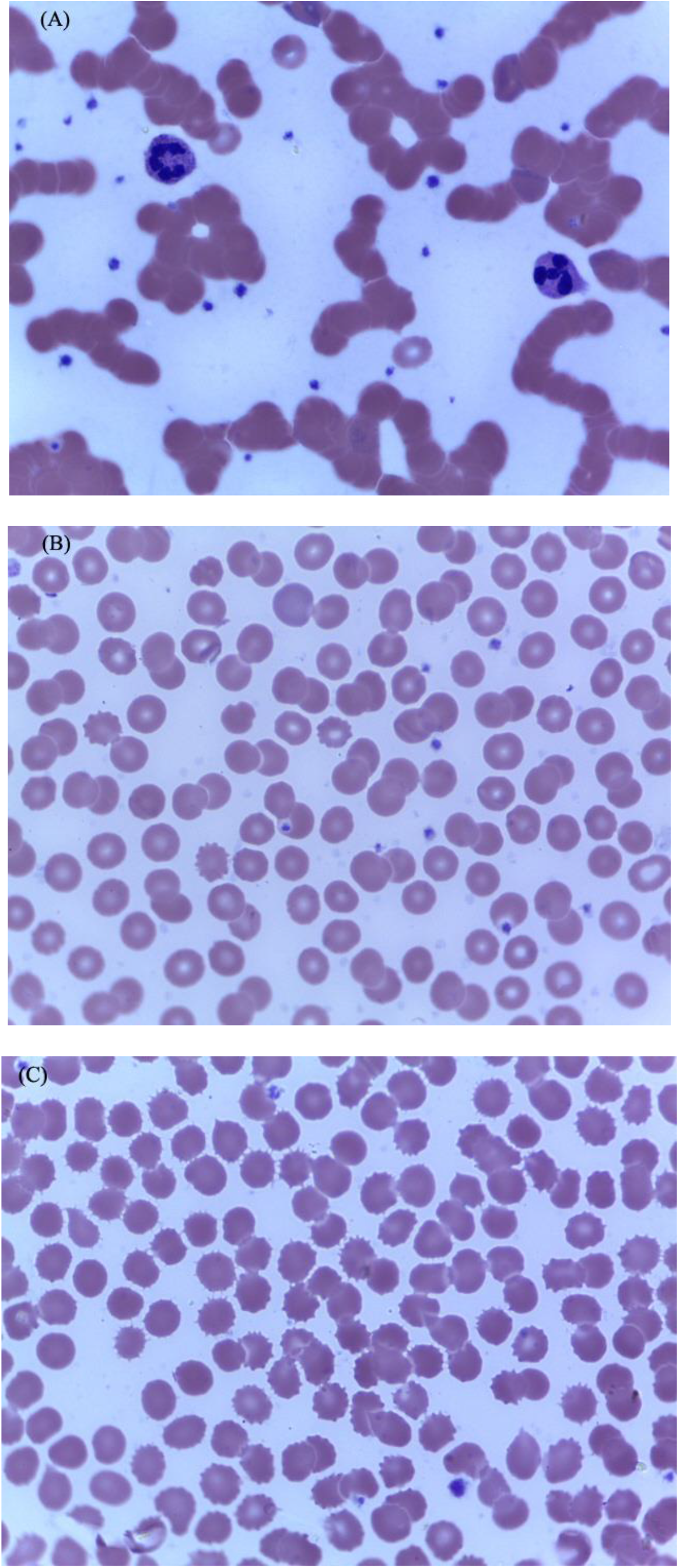
Red blood cells injury and hypersegmented neutrophils in peripheral blood films from COVID-19 patients at the hospital admission. A, Hypersegmented neutrophils and rouleaux formation. B, Spherocytes with the disappearance of biconcave disk. C, Echinocytes showing spiky projections.

## Data Availability

All the data present in the current study are available at the University of Bejaia

